# Enteric viruses isolates and rotavirus genotypes during the seasonal surge of acute watery childhood diarrhoea in South-Eastern Nigeria

**DOI:** 10.1101/2022.03.25.22272944

**Authors:** Akinwale Michale Efunshile, Chukwuemeka Nwangwu, Bethrand Amechi Ngwu

## Abstract

Diarrhoea remains one of the top three causes of death in Africa. However, data on the molecular epidemiology of enteric viruses in Nigeria is rare.

Two hundred and forty three infants and children below the age of 5 years with acute watery diarrhoea were evaluated for the presence of enteric viruses in stool by real-time PCR (rtPCR) during the dry months of December to April 2017, which correspond to diarrhoea season in South-East Nigeria. At least one viral pathogen was detected in 95.1% (231/243) of the study population. Rotavirus, 231(95.1%) was the most common followed by adenovirus, 103(42.4%) and enterovirus 32(13.2%). Other viruses seen in the stool samples include astrovirus 7.3 % (11/151), parechovirus 3.5 % (4/113), sapovirus, 2.8 % (4/145), bocavirus 6.8% (5/73) and human coronavirus 13.9% (10/73). Norovirus and hepatitis A and E viruses were not detected. Children that consumed factory packaged water had the lowest prevalence of rotavirus infection (p=0.044). A significant association between the viral pathogen and home treatment of drinking water or exclusive breastfeeding was not observed. Randomly selected 46 samples were genotyped for rotavirus, which showed that G3P[6] (39.1%) was the most common, followed by G1P[8] (15.2%), G9P[8] (13.0%), G12P[8] (6.5%), G9P[6]P[8] (2.2%), and G12P[6] (2.2%).

This was the highest rotavirus prevalence in any published African study, and may be a reflection of inadequate water sanitation/hygiene in the study area, a peculiar epidemiological situation and/or the sensitivity of the diagnostic method used.

The present study suggests that the burden of rotavirus is bigger than previously reported; and that morbidity can be greatly reduced if rotavirus vaccine is included in Nigerian national immunization policy.

## Introduction

Diarrhoea caused an estimate of 1.3 million deaths globally in 2015 and it was reported to be the 4^th^ leading cause of death among children less than 5 years old [1].

Diarrhoea was reported to have killed more than 500.000 children under 5 years in 2015 alone, with Asia and Sub-Saharan Africa recording the highest number of incidence and deaths [2]. Although, deaths from diarrhoea and other diseases declined globally between 2010 and 2015, the reduction rate was not remarkable in Nigeria and many other Sub-Sahara Africa countries as a result of inadequate water, poor sanitation and hygiene; as well as predisposition from malnutrition [2]. Rotavirus, cryptosporidium and shigella *spp* were the major agents which accounted for more than 50% of diarrhoea deaths among children below the age of 5 years in 2015 [1]. In its comprehensive assessment, the Global Burden of Diseases in 2015 also listed enteric adenovirus and noroviruses as other viral agents that significantly contributed to diarrhoea deaths [1]. Though, the burden of specific viral agent depends on the study region and the season. Because rotavirus is the commonest diarrhoea agent worldwide, the incidence and mortality of its associated diarrhoea were shown to have significantly declined in countries where rotavirus vaccines were introduced [3]. The WHO therefore recommends that rotavirus vaccine is included in routine immunization policy, especially in developing countries [4]. Rotavirus gastroenteritis is more seasonal in temperate regions compared to the tropical areas where it occurs more or less all the year round. And this difference was found to be associated with socio-economic factors rather than being an effect of climate [5]. A systematic review of rotavirus diarrhoea in several African countries concluded that the disease usually peak during the dry period of the year in most countries [6]. In Asia, where diarrhoea burden is similar to what obtains in Africa, findings also showed that diarrhoea occurs all the year round, and that 45% of diarrhoea in children below the age of 5 years was attributed to rotavirus [7]. Co-infection between rotavirus and other viral diarrhoea agents have also been reported. For instance, rotavirus (26.6%) was found to be more common among Moroccan diarrhoea children, while mixed infection with norovirus was found in 2.7% of the cases [8]. Another diarrhoea study reported rotavirus and norovirus in a cohort of 50 diarrheic South-West Nigerian children with a prevalence of 34.5% and 25.5% respectively, while co-infection with both viruses was seen in 9.1% of cases [9]. Three enteric viruses, rotavirus (28.3%), Adenovirus (19.3%), and norovirus (3.6%) were also reported among diarrhoea children in South-South region of Nigeria with co-infections by 2 or 3 viruses in 7.6% cases [10]. A recent report evaluating diarrhoea agents over a 12 months period in Nigeria showed that at least one was identified in 58.3% of cases with co-infections involving norovirus or astrovirus or aichivirus in some cases [11].

Countries with the highest rotavirus burden including Nigeria are yet to implement the WHO vaccine policy despite its proven efficacy [4,12]. Changes in the aetiology and epidemiology of viral agents of diarrhoea were reported following the introduction of rotavirus vaccines [13, 14]. This underlines the importance of continuous surveillance of enteric viral agents to enable effective planning for future control efforts. Such surveillance will also help to bring the public health impact and effectiveness of the vaccines to the forefront, thereby encouraging policy implementation in countries that are yet to comply. The present study aimed at evaluating enteric viral agents associated with acute diarrhoea in the study area and to provide baseline epidemiologic data ahead of the national rotavirus vaccine implementation. Additionally, the relationship between feeding practices and sources of drinking water and enteric viruses was evaluated.

## Methods

### Study design

This is a cross sectional study primarily designed to identify the viral agents of acute diarrhoea in infants and children below 5 years of age. Inclusion criteria were passage of watery stool three or more times within the last 24 hours, age of 5 years or less and willingness to participate in the study. The Ethical Committee of the Federal Teaching Hospital Abakaliki, Nigeria, approved the study. Written informed consent were obtained from the parents/guardian of the children after the purpose of the study were translated to them in local dialects.

### Study Area

The study was carried out at University Teaching Hospitals (Ebonyi and Enugu) in the South-Eastern Region of Nigeria between latitudes 04^0^30’N and 07^0^30’N and longitudes 06^0^45’E and 08^0^45’E [15], during the peak diarrhoea period, in the months of December 2016 to April 2017.

### Data collection and management

Questionnaires were administered to the parents/guardian of the participants to collect data regarding age, frequency and duration of diarrhoea, risk factors and other relevant information. This information were subsequently transferred into excel sheets for analysis.

### Specimen management and preliminary examination

About 5 ml of watery stool were collected from the participants into wide mouth bottles. Inactivated aliquots were shipped to the Institute of Virology, Leipzig University, Germany.

### Virus PCR

Total nucleic acids was extracted from 10 % stool suspensions in PBS by MagNA Pure 96 system (Roche Applied Science, Manheim, Germany), aliquoted, and stored at -80°C.

Aliquots were subsequently screened by real-time (RT)-PCRs for presence of viral genome of species A rotavirus, norovirus (genogroup I and II), sapovirus (genogroup I, II, IV and V), adenovirus (group A to F), parechovirus, astrovirus (HastV 1–8), and hepatitis E virus and coronavirus [16-24]. Primers and probes used for detection of enterovirus (EV) species A to D (including coxackieviruses, echoviruses, enterovirus and poliovirus), human bocavirus and hepatitis A virus (HAV) targeted the 5’NTR-regions of the viral genome: EV-1s (5’-GGTGCGAAGAGTCT ATTGAGCTA-3’), EV-1as (5’-ACCCAAAGTAGTCGGTTCC-3’), EV-probe (5’-6FAM-TGAATGCGGCTAATCCTAACTGCG-DB-3’), hBoV_5UTR_Q_s1 (5’-CCTATATAARCCGATGCACT), hBoV_5UTR_Q_s2 (5’-CCTATATAAGCTGCTGCACT), hBOV_NS1_Q_as1 (5’-AGGAGCAGAAAAGGCCATA), hBoV_NS1_Q_as2 (5’-AGGRGGATTGAAAGCCATA), hBoV_Q-TaqFAM (5’-6FAM-TCCGGCGAGTGAACATCTCT—BBQ), HAV-5UTR-Qs (5’AATATTAATTCCTGCAG GTTCAG-3’), HAV-5UTR-Qas (5’AGCCAAGTTAACACTGCAAG-3’), and HAV-5UTR QFAM (5’6FAM-CAATTTAGACTCCTACAGCTCCATGCTA-BBQ-3’).

### Rotavirus G and P typing

Randomly selected 46 samples tested by quantitative real time PCR for rotavirus were sequenced as previously described using published methodology [25,26]. The G and P type-specific amplification of VP7 and VP4 fragments was performed using QIAgen One-step RT-PCR kit (QIAgen, Hilden, Germany). For the first run, primers used were VP7-F and VP7-R for the VP7 gene while VP4-F and VP4-R were used for the VP4 gene. A second semi-nested multiplex PCR was done using G1-, G2-, G3-, G4-, G8-,G9-, G10- and G12-specific sense primers for G types, while P-typing included P[4]-, P[6]-, P[8]-, P[9]-, P[10]-, and P[11]-specific antisense primers together with VP7-R and VP4-F, respectively.

Gel purified amplicons (Wizard SV Gel and PCR Clean-Up System; Promega, Mannheim, Germany) were partially sequenced (approximately 600 bp each) using BigDye Terminator v1.1 Cycle Sequencing kit (PE Applied Biosystems, Foster City, CA) on an ABI Prism 310 Genetic Analyzer. Nucleotide sequences read from the chromatograms were aligned.

Sequencing of the r amplicons was done for the 46 randomly selected rotavirus isolates.

### Statistical analysis

Data were analysed with the freely available GNU PSPP Statistical Analysis Software version 1.0.1-g818227 [27]. Descriptive statistics was used to compare the various variables, while Chi-square tests were used to derive significant associations which was set at p-value <0.05.

## Results

For the present investigations, stool samples obtained from 243 children were available. Most of the children, 71.2% (173/243) were below the age of 2 years, and 65% (158/243) were males (table 1).

**Table 1.** Age and sex distribution of participants.

|  | Gender (N (%)) |  | Total |
| --- | --- | --- | --- |
|  | Male | Female |  |
| Age group |  |  |  |
| <2 | 111 (64.2) | 62 (35.8) | 173 (100) |
| 2-3 | 40 (64.5) | 22 (35.5) | 62 (100) |
| >3 | 7 (87.5) | 1 (12.5) | 8 (100) |
| Total | 158 (65.2) | 85 (34.8) | 243 (100) |

Eleven enteric viruses were tested. Not all samples were tested for each of the viruses (table 2). Ninety-four (38.7%) of the adenovirus positive cases were also co-infected with *Rotavirus* while triple infection between rotavirus, adenovirus and enterovirus was observed in 7.0% of the cases (table 2).

**Table 2.** Enteric virus distribution in the study population.

| Enteric viruses | Number of samples tested | Prevalence, N (%) |
| --- | --- | --- |
| Rotavirus | 243 | 231 (95.1) |
| Adenovirus | 243 | 103 (42.4) |
| Enterovirus | 243 | 32(13.2) |
| Astrovirus | 151 | 11(7.3) |
| Sapovirus | 150 | 3(2.0) |
| Norovirus | 145 | 0(0.0) |
| Parechovirus | 113 | 4 (3.5) |
| Bocavirus | 73 | 5(6.8) |
| Human Coronavirus | 73 | 10(13.7) |
| Hepatitis A virus | 51 | 0(0) |
| Hepatitis E virus | 51 | 0(0) |
| Rotavirus +Adenovirus | 243 | 94(38.7) |
| Rotavirus +Enterovirus | 243 | 30(12.3) |
| Rotavirus +Astrovirus<sup>#</sup> | 151 | 7(4.6) |
| Adenovirus + Enterovirus | 243 | 24(9.9) |
| Rotavirus +Adenovirus + Enterovirus | 243 | 17(7.0) |
| Enterovirus +Sapovirus<sup>#</sup> | 150 | 1(0.7) |
| Parechovirus<sup>#</sup> +Adenovirus | 113 | 1(0.9) |

N; number of positives <sup>#</sup> See discussion

There was no significant association between ages or sex and any of the 3 most commonly encountered enteric viruses (table 3).

**Table 3.**
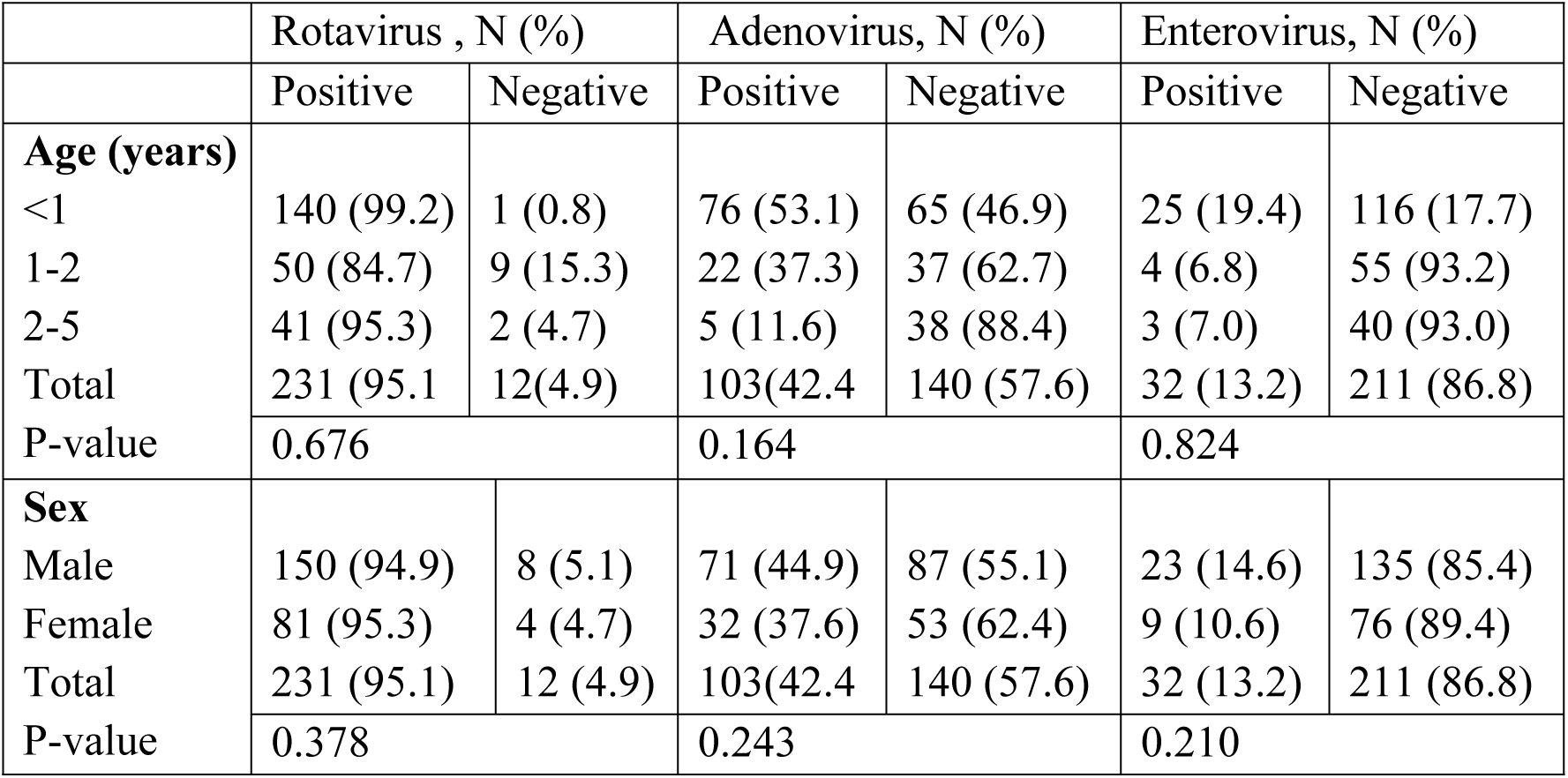
Age and sex distribution of *Rotavirus, Adenovirus* and *Enterovirus* among diarrheic children below the age of 5 years.

The present study did not establish a significant association between the three common enteric viruses (i.e. rotavirus, enterovirus and adenovirus) and past history of exclusive breastfeeding or the current feeding practice of the children (table 4).

**Table 4.** Rotavirus, adenovirus and enterovirus prevalence in relation to feeding practices in diarrheic children below the age of 5 years.

|  | Rotavirus |  | Adenovirus |  | Enterovirus |  |
| --- | --- | --- | --- | --- | --- | --- |
|  | Pos, N (%) | Neg , N (%) | Pos, N (%) | Neg, N (%) | Pos, N (%) | Neg, N (%) |
| **EBF history** |  |  |  |  |  |  |
| Yes | 102 (95.3) | 5 (4.7) | 47 (43.9) | 60 (56.1) | 37 (34.6) | 70 (65.4) |
| No | 129 (95.6) | 6 (4.4) | 61 (45.2) | 74 (54.8) | 24 (17.8) | 111 (82.2) |
| Total | 231(95.1) | 12(4.9) | 103(42.4) | 140 (57.6) | 32 (13.2) | 211 (86.8) |
| P-value | 0.378 |  | 0.055 |  | 0.496 |  |
| **Current Feeding** |  |  |  |  |  |  |
| EBF | 16 (100) | 0(0) | 7 (43.8) | 9 (65.2) | 2 (12.5) | 14 (87.5) |
| Complimentary | 195 (95.1) | 10 (4.9) | 87 (44.5) | 118 (57.6) | 28 (13.7) | 177 (86.3) |
| PBF | 19 (95) | 1 (5) | 8 (40.0) | 12 (60.0) | 1 (5.0) | 19 (95.0) |
| Supplementary | 1 (50) | 1 (50) | 1 (50.0) | 1 (50.0) | 1 (50.0) | 1 (50.0) |
| Total<br>P-value | 231(95.1) | 12(4.9) | 103(42.4) | 140 (57.6) | 32 (13.2) | 211 (86.8) |
|  | 0.85 |  | 0.981 |  | 0.153 |  |

Key-Pos= Positive, Neg= Negative, EBF= Exclusive breastfeeding, PBF = Predominant breastfeeding

But there was a significant association between the sources of drinking water and prevalence of rotavirus infection (P=0.044), as children that consumed factory packaged water had the lowest prevalence of rotavirus infection. But this association was not significant for adenovirus and enterovirus (table 5). Approximately half of the children consumed factory packaged water from plastic bottles. An association between home treatments of drinking water with any of the viruses was not observed, though the majority of children consumed water without any form of home treatment (table 5).

**Table 5.** Rotavirus, adenovirus and enterovirus relationship with sources of drinking water and treatment practices among children below the age of 5 years.

|  | Rotavirus |  | Adenovirus |  | Enterovirus |  |
| --- | --- | --- | --- | --- | --- | --- |
|  | Pos, N (%) | Neg, N (%) | Pos, N (%) | Neg, N (%) | Pos, N (%) | Neg, N (%) |
| **Source of drinking water** |  |  |  |  |  |  |
| Cart vendor | 66 (100) | 0 (0) | 30 (45.5) | 36 (55.5) | 8 (14.0) | 49 (86.0) |
| Borehole | 13 (92.9) | 1 (7.1) | 8 (57.1) | 6 (42.9) | 6 (42.9) | 8 (57.1) |
| Dug well | 4 (100) | 0 (0) | 1 (15.0) | 3 (75.0) | 2 (50.0) | 2 (50.0) |
| Bottled water | 116 (96.7) | 4 (3.3) | 57 (43.2) | 75 (56.8) | 16 (13.5) | 102 (86.5) |
| Total<sup>#</sup> | 199 (97.5) | 5 (2.5) | 96(44.4) | 120 (55.6) | 32(16.6) | 161(83.4) |
| P-value | 0.044* |  | 0.691 |  | 0.097 |  |
| **Home water treatment** |  |  |  |  |  |  |
| None | 165 (97.1) | 5 (2.9) | 75 (41.2) | 107 (58.8) | 30 (18.6) | 130 (81.4) |
| Boiling | 30 (100) | 0 (0) | 17 (56.7) | 13 (43.3) | 2 (6.7) | 28 (93.3) |
| Filtration | 1 (100) | 0 (0) | 1 (100.0) | 0 (0) | 0 (0) | 1 (100) |
| Chemical (Chlorination) | 3 (100) | 0 (0) | 3 (100.0) | 0 (0) | 0 (0) | 2 (100) |
| Total | 199 (97.5) | 5 (2.5) | 96(44.4) | 120 (55.6) | 32 (16.6) | 161 (83.4) |
| P-value | 0.338 |  | 0.051 |  | 0.155 |  |

Key-Pos= Positive, Neg= Negative, \*= Significant value, # Not all children drink water because of exclusive breastfeeding

There was no significant difference between rotavirus infections in the children across the two cities that were studied. Adenovirus was significantly more common among the children from Enugu (P=0.001), (table 6).

**Table 6.** Rotavirus, adenovirus and enterovirus distribution across the study areas.

| Study sites | Rotavirus, N (%) |  | Adenovirus, N (%) |  | Enterovirus, N (%) |  |
| --- | --- | --- | --- | --- | --- | --- |
|  | Pos | Neg | Pos | Neg | Pos | Neg |
| Enugu | 161 (95.3) | 8 (4.7) | 86 (50.8) | 83 (49.2) | 31 (18.3) | 138 (81.7) |
| Abakaliki | 70 (94.5) | 4 (5.5) | 17 (22.9) | 57 (77.1) | 1 (1.4) | 73 (98.6) |
| Total | 231(95.1) | 12(4.9) | 103(42.4) | 140(57.6) | 32 (13.2) | 211 (86.8) |
| P-value | 0.418 |  | 0.001* |  | 0.048* |  |

Key-Pos= Positive, Neg= Negative, \*= Significant value

The genotypes of the 46 randomly selected rotavirus isolates were described in the table 7. Genotype G3P[6] was the predominant genotype, 18(39.1) followed by G1P[8], 7(15.2).

**Table 7 :** Distribution of G and P genotype combination of RVA strains detected among gastroenteritis children below 5years old.

| Rotavirus genotype |  |  |  |  |  |
| --- | --- | --- | --- | --- | --- |
|  | No. (%) of strains |  |  |  | Total |
|  | P[6] | P[8] | P[6]/P[8] | P n/t |  |
| G1 | 0 | 7(15.2) | 0 | 0 | 7(15.2) |
| G3 | 18(39.1) | 0 | 0 | 0 | 18(39.1) |
| G9 | 0 | 6(13.0) | 1(2.2) | 0 | 7(15.2) |
| G12 | 1(2.2) | 3(6.5) | 2(4.3) | 0 | 6(13.0) |
| G1/G9 (mixed) | 0 | 0 | 1(2.2) | 0 | 1(2.2) |
| G3/G12 (mixed) | 1(2.2) | 0 | 2(4.3) | 0 | 3(6.5) |
| G9/G12 (mixed) | 0 | 1(2.2) | 1(2.2) | 0 | 2(4.3) |
| G n/t | 2(4.3) | 0 | 0 | 0 | 2(4.3) |
| Total | 22(47.8) | 17(14.0) | 7(15.2) | 0 | 46(100) |

n/t= not typeable

## Discussion

The prevalence of rotavirus observed in our study (95.1%) was far higher than the average report among African children, which is in the range of 25 to 40% [6]. It was also higher than the WHO rotavirus surveillance data which reported a mean prevalence of 32% among 37 countries including Nigeria [28]. The High prevalence might be due to differences in water supply and hygiene practices between our study area and the other regions with lower prevalence. For instance, a recent review showed that packaged water in developing countries was 4.6-13.6 times more likely to contain bacteria indicator of faecal contamination compared to developed countries [29]. Sizable numbers of the children also drank from cart water vendors, while most of the water consumed was not subjected to any form of home treatment. Differences in the prevalence of rotavirus may also depend on the diagnostic method used and the season of the year when the study was conducted. It is likely that the present study would have reported a lower prevalence if ELISA had been used, because a study by the research group of Adah reported a rotavirus prevalence of 11.8% and 88.2% from the same stool samples, when ELISA and RT-PCR methods were used respectively [30]. Similar studies from our study area reported a lower rotavirus prevalence of 46% and 56% in 2 separate studies carried out over a period of 5 and 2 years respectively using ELISA test methods [31, 33]. But another Nigerian study that used RT-PCR also reported lower rotavirus prevalence of 48%. This might be related to the differences in the study regions and study period [33].

Lack of significant association between enteric viruses and exclusive breastfeeding in the present study seems unpredictable. This is possibly because we did not evaluate non-diarrhoeic children as negative controls, which of course is beyond the scope of our study. But there are studies which seem to agree with our findings, for instance, the research group of Clemens showed that breastfeeding does not reduce incidence of diarrhoea, it only reduces the severity [34]. Golding *et al* also concluded in their research that breastfeeding only protect against diarrhoea of non-viral aetiologies [35].

The 42.4% prevalence of Adenovirus in our study was higher that the result from a study from North-Western Nigeria [36] which was 22.3%. The difference may also be because we used RT-PCR technique in our study which is more sensitive than the rapid kits used by the other study. Unlike our findings, another study from Nigeria reported higher prevalence of astrovirus (40.4%) [37], and this might be due to regional differences in viral diarrhoea agents. Absence of norovirus in our study was contrary to findings from other African studies. Norovirus prevalence of 25.5% was once reported in South-western region of Nigeria while a review of 19 studies from Sub-Sahara Africa showed that the prevalence ranged from 4.6% to 32.4% [38]. The season of our study was probably responsible. There is a need to further investigate the epidemiology of this virus in South-East Nigeria in a larger cohort of diarrheic children, with specimen collection throughout the year. Studies relating to sapovirus are very rare in Nigeria. A similar study in 2 areas in Kenya also reported low prevalence of 4% and 6% at both areas [39]. This supports the impression that this enteric virus played less significant role in our current study. Like sapovirus, this is the first report of parechovirus study in Nigeria. Two of the three samples that were positive for this virus were negative for all other enteric viruses tested, and they also had the cycle threshold (C_T_) of the RT-PCR below 30 (Ct data not shown), suggesting significantly high viral load and probability of being responsible for the diarrhoea cases. But it had been reported that the types, rather than viral load of parechovirus is more important in clinical settings. Infections with type 3 HPeV is said to be clinically relevant when detected, regardless of the viral load [40]. Co-infection of 2 or 3 enteric viruses observed in our study was likely due to the fact that they share the same faecal-oral transmission pathway, as a result of inadequate water, sanitation and hygiene. The present result of co-infection with astrovirus, sapovirus and parechovirus also called for interpretation with caution since only a subset of the stool samples were tested for these particular viruses. Further studies will need to simultaneously test larger sample size to get a more accurate co-infection rate. The relatively lower C_T_ of *rotavirus* compared to other viruses is a reflection of its higher viral load [41], which may suggest its relative importance in the cases with co-infections.

From this study, genotype G3P[6] was the commonest genotype observed. This supports the earlier suggestion that the high prevalence of the RTV reported in this study could be largely due to ingestion of contaminated water since type G3 is known to be conserved in human without zoonotic transmission from animals.^42^ Some authors have observed a significant correlation between G3 genotype and poor sanitation and infection control, and low rotavirus vaccine coverage [42-44]. Although most recent studies in Nigeria, reported other rotavirus G types as the most common. For instance in studies done by Japhet (2017) and Motayo (2018), the prevalent genotypes were G12 and G1 respectively [43, 44]. The two studies were done throughout the year in the Western part of Nigeria. Apart from the differences in the study period, the study areas were different from ours with respect to social, cultural and economical determinants. These factors were known to determine the spread of rotavirus genotype [45], hence there is need for national surveillance and characterization of rotavirus in Nigeria for accurate data to better influence policy, monitoring and evaluation of interventions. In a six year (2011-2016) rotavirus surveillance study done at Enugu by Tagbo (2019), G3P [6] was the most common genotype reported in 2016, which is the part of the period our study was conducted [46]. This also affirms the yearly and seasonal variation of rotavirus strains as wildly reported [45]. In the same study by Tagbo, the cumulative prevalence of rotavirus genotypes throughout the six year study period was G12, G1 and G3 accounting for 27.6%, 21.0% and 16.3% respectively [46]. Therefore rotavirus strains distribution and prevalence is determined by a lot of factors and study design.

## Conclusion

The present study suggests that the burden of rotavirus in Nigeria was likely underestimated by previous studies that used non-molecular epidemiologic tool. It also underscores the need to properly investigate the accurate epidemiology of enteropathogenic viruses at a national level so that attention of policy makers and other healthcare stakeholders can urgently be drawn to the need for prompt implementation of rotavirus vaccine as part of the national immunization policy.

## Data Availability

Sample of the questionnaire used for data collection will be made available

## Limitation of the study

The study did not rule out all other possible cause of diarrhoea including bacteria and parasites among the study participants, so it was difficult to establish a causal relationship between the viruses and the diarrhoea.

## Author’s contributions

EAM, NC conceived the project idea and developed the proposal. EAM, NC carried out sample and data collection, NC went to Germany to analyze the samples, EAM, NC, NBAF wrote the manuscript, EAM, BAF carried out the statistics analysis. All the authors edited and approved final version of the manuscript

## Acknowledgment

We sincerely thank the Institute of Virology, Leipzig University, Germany for the technical assistance for the molecular diagnostics

